# The implementation of a virtual ward using digital solutions informing community clinicians in early supported discharge of patients with SARS-Cov2 respiratory symptoms from an acute hospital setting

**DOI:** 10.1101/2021.03.29.21254548

**Authors:** J Swift, Z Harris, A Woodward, NI O’Kelly, C Barker, S Ghosh

## Abstract

**Objectives:** To assess the short run successes and challenges of the implementation of a digitally supported accelerated acute hospital discharge scheme for patients admitted with Covid-19.

**Design:** Analysis of the safety, resource use and health outcomes within the virtual service for the first 65 patients that have been discharged from a virtual respiratory ward.

**Setting:** Community based intervention using digital technology and a multi-disciplinary team of specialist clinicians to monitor patients at home.

**Participants:** 65 patients discharged from hospital followed until discharge from the virtual ward.

**Results:** 24.6% of 65 patients had symptoms that were coded red (urgent response required) in CliniTouch Vie in the first day after hospital discharge falling to 7.7% on their final day on the virtual ward; p=0.049. Reductions in red days decreased significantly over time, from 33.8% of patients in their first three days to 10.8% in their final three days; all patients p=0.002. Four patients were re-admitted to hospital, all for clotting disorders. There was one death within this group, which following senior clinical review was deemed to be unrelated to infection with Covid-19.

The most important gain for Glenfield hospital was in expediting the rapid discharge of patients admitted with Covid-19 into a supported environment and the freeing up of beds. On 15^th^ January, 48% of beds were taken up with patients admitted with Covid-19 symptoms.

In November 2020, immediately prior to the launch of the virtual ward, the mean length of stay for patients who did not access high dependency care or oxygen was 5.5 (+/-1.3) days. The mean length of stay in patients discharged into the virtual ward thereafter was 3.3 (+/-0.4) days; relative reduction, 40.3% (p<0.001).

The cost of care provision in the virtual ward was 8,662 UK Pounds in total and 133.26 UK pounds per patient. The estimated overall savings were 68,052 UK Pounds and the mean saving per patient was estimated at £1,047 UK Pounds.

**Conclusions:** The virtual ward appeared to assist with earlier discharges, had a low rate of clinically necessary re-admissions, the safety of patients was not compromised and whilst cost savings were not the primary objective, it seemed to also reduce overall resource use and costs.

## Objective of project / paper

This paper reports on the deployment and early clinical and economic outcomes of a digital pathway to allow patients admitted with SARS-CoV2 to be discharged from acute hospital care and monitored within a community setting by a specialised community respiratory team. Although this is an early evaluation, it was felt that sharing the description of the service and early outcomes at a time when healthcare systems around the world have been facing unprecedented challenges was important.

## Background

COVID-19 is caused by the coronavirus, SARS-CoV 2. The disease shares similarities with both SARS-CoV and MERS-CoV in causing a fulminant immunopathological response, leading to Acute Respiratory Distress Syndrome (ARDS) in a proportion of patients. The pathology, transmissibility, impact upon hospital capacity and policy responses have been widely reported in this journal and elsewhere.

During the first wave there was concern that health systems could be overwhelmed. It was expected this change in the model of care would result in a release of pressure on the acute setting amongst other potential benefits to patients and healthcare professionals.

Leicester, Leicestershire and Rutland (LLR), like the rest of the United Kingdom has experienced two significant surges in Covid-19 infections and hospital admissions. This has caused unprecedented issues with the provision of hospital-based care within LLR and the University Hospitals Leicester NHS Trust (UHL) declaring a highest alert in the middle of December 2020.

During the first wave of the pandemic between May and June in 2020, a digital pathway for assisted discharge for patients with Covid-19 was devised by clinicians from UHL, the community specialist respiratory team of Leicestershire Partnership Trust (LPT) and Spirit Digital. The clinical algorithms used within the Covid-19 digital platform were devised and reviewed by two independent respiratory consultants.

The digital platform was deployed in November 2020 when LLR experienced the second wave of the pandemic. Its onset coupled with annual capacity issues experienced by acute hospitals during winter created severe challenges in the delivery of acute care within LLR. This was further compounded by the impact that Covid-19 had on the workforce with a number of clinicians either suffering themselves from Covid-19 and self-isolating or being shielded from direct patient contact because of potential vulnerabilities.

CliniTouch Vie was already in use in LLR to digitally support the distal management of patients with respiratory conditions. Locally there was clinical recognition of the bifurcation of patients into two separate cohorts during the first wave, those likely to recover quickly and those expected to deteriorate further gave a plausible hypothesis to the latter cohort being eligible to access an early supported discharge intervention.

The challenge was to find a solution to create greater flow in the system by enabling patients at the early stages of recovery from a Covid-19 infection to be discharged safely into the community. It was recognised that this required a system-wide approach with acute hospital services working closely with community provision to ensure patients were monitored following their discharge. A model was needed to be flexible enough to respond to further potential surges in the incidence of covid-19 and to also allow for the impact on local capacity exacerbated by annual winter pressures.

### Service Description

The virtual ward was designed to avert a potential crisis in patient flow and system capacity caused by a likely excess of patient admissions related to the Covid-19 pandemic should there have been a second wave. The supported discharge and enhanced monitoring service provided remote support and follow-up for patients admitted to hospital with a clinical diagnosis of Covid-19 and at risk of deterioration after discharge. It provided care for patients in LLR, in a partnership between LPT, UHL and Spirit Digital. CliniTouch Vie is a digital platform that comprises both a patient and clinical portal. Clinical data is entered by the patient and uploads to a clinical dashboard, which prioritises patients depending on their responses to clinical questions and observations. The platform also allows for patients and clinicians to connect via messaging or by embedded video consulting features. The platform can be accessed via any SMART phone or tablet (Android or IOS) or any computer (Windows or Apple operating systems).

### Discharge / referral procedures and criteria

The specialist respiratory department at UHL is located within Glenfield Hospital. Patients were identified for the service following a senior medical review. The preference was for patients to use their own device, either a smartphone, tablet or laptop. However, if patients did not have access to a suitable device, a smartphone was provided free of charge by Spirit Digital.

At the time of implementation, due to the lack of a validated deterioration risk scoring tool for Covid-19 patients, a clinical decision aid was formulated by the respiratory team at UHL to assess suitability for discharge and appropriate safety netting can be found in table 1 below.

**Table 1:** Clinical referral criteria and risk categorisation

|  |  |  |
| --- | --- | --- |
| - Medically fit for supported discharge as decided by their designated consultant<br>- Able to cope at home<br>- For full active treatment<br>- In possession of a smartphone/tablet/laptop<br>- **Two or more** risk factors for increased safety netting, from the list below: |  |  |
| **Risk factors for increased safety netting:** |  |  |
| < 10 days symptoms onset | RR 20-22 | Extremely vulnerable group<br>(as per PHE advice) |
| Oxygen saturations 92-94% | Lives alone |  |

### Onboarding process

Once patients were identified as suitable to be discharged and monitored within their own homes, a structured onboarding process was initiated by UHL ward and in-reaching LPT staff (Appendix 2).

## Methods

### Overall Summary

The analysis is primarily narrative in nature and describes the early results of the implementation of the virtual ward programme. Whilst statistical methods have been employed to determine the validity of the initial findings, many endpoints were not pre-specified, no estimate of effects pre-determined, no power calculations made and all data analysed were observational. The data were open to bias and confounding and the results should be interpreted accordingly. This reflects the environment within which the ward was introduced as a pre-planned, yet emergency response to circumstances.

### Key Outcomes

The following were considered to be the most important endpoints to be evaluated; a priori in bold and post hoc in plain text.

- **The overall safety of the virtual ward; i.e. the number and percentage of re-admissions and the number and distribution of red days in the virtual ward**
- **The hospital length of stay in the population that accessed the virtual ward** and to establish if bed days were reduced post the introduction of the virtual ward
- **The costs associated with the virtual ward** and estimated impact upon overall resource use

### Analysis of patient health status

Every patient accessing the virtual ward had their daily risk; red, amber, green (RAG) rating recorded. The numbers of patients in the virtual ward and their RAG status were explored over time graphically and statistically. Student’s t-tests (paired and two tailed) were conducted to compare patients’ scores in their first day, first three days, versus their last day, last three days. These data were extracted from the CliniTouch database and analysed in Microsoft Excel 365 Data Analysis pack. All re-admissions were reported with the cause of re-admission identified and case explored by senior clinicians.

### Patients’ length of stay

Treatment regimes in the first and second wave of the pandemic had altered, as clinical knowledge improved. The patients that accessed the virtual ward were compared to the entire cohort of patients who did not require either oxygen or intensive monitoring prior to the introduction of the virtual ward in November 2020 during the second wave, i.e. broadly comparable patients and treatments. Of the 66 patients admitted into the ward, there were data for 65, one patient was re-admitted prior to inputting any data. This admission and costs are included in the narrative, but the denominator has been kept as 65 to maintain consistency.

### Resource use associated with the intervention

Clinical resource use information on the virtual Covid-19 ward was provided by LPT and the costs of clinician time attached to the intervention were sourced from The University of Kent’s PSSRU dataset^1^. The number of interventions were accessed from the CliniTouch Vie database. The cost of a bed day in the Glenfield Hospital respiratory wards and individual lengths of stay for patients admitted into the virtual ward and the mean plus confidence interval for similar patients (no intensive or high dependency care or oxygen administration) admitted and discharged throughout the month of November and immediately prior to the introduction of the virtual ward in Glenfield Hospital were sourced from the business team within the UHL Finance Directorate in cooperation with the Clinical Commissioning Groups for LLR^2^.

#### All further analyses were based upon these datasets

No sample sizes were pre-specified for this analysis, this was an early analysis designed to explore the impact of the intervention.

### Patient demographics

Participant ages ranged from 21.5 to 87.4years with a mean and median age of 56 and 58years. 39% of patients were female and no ethnicity data were retained.

## Results

The number of patients admitted to the LPT supported discharge virtual ward as of 31/1/2021 was 66. The results are presented for the 65 patients that inputted data into CliniTouch Vie. The number of patients re-admitted was 4 (6.2%) and included the patient for whom there was no data.

The number of patient red alerts on day one was 16 (24.6%) and in the first three days was 23 (33.8%). The number of patients with a red alert on their final day was five (7.7%) and in the last three days was seven (10.8%); first versus last day, relative reduction 56.3%, p=0.049. The relative reduction for the first three days versus the last three days; 68.2%, p=0.002.

The absolute number of RAG rating scores over time are shown in Figure 1 beneath for the 65 participants in the cohort.

**Figure 1.**
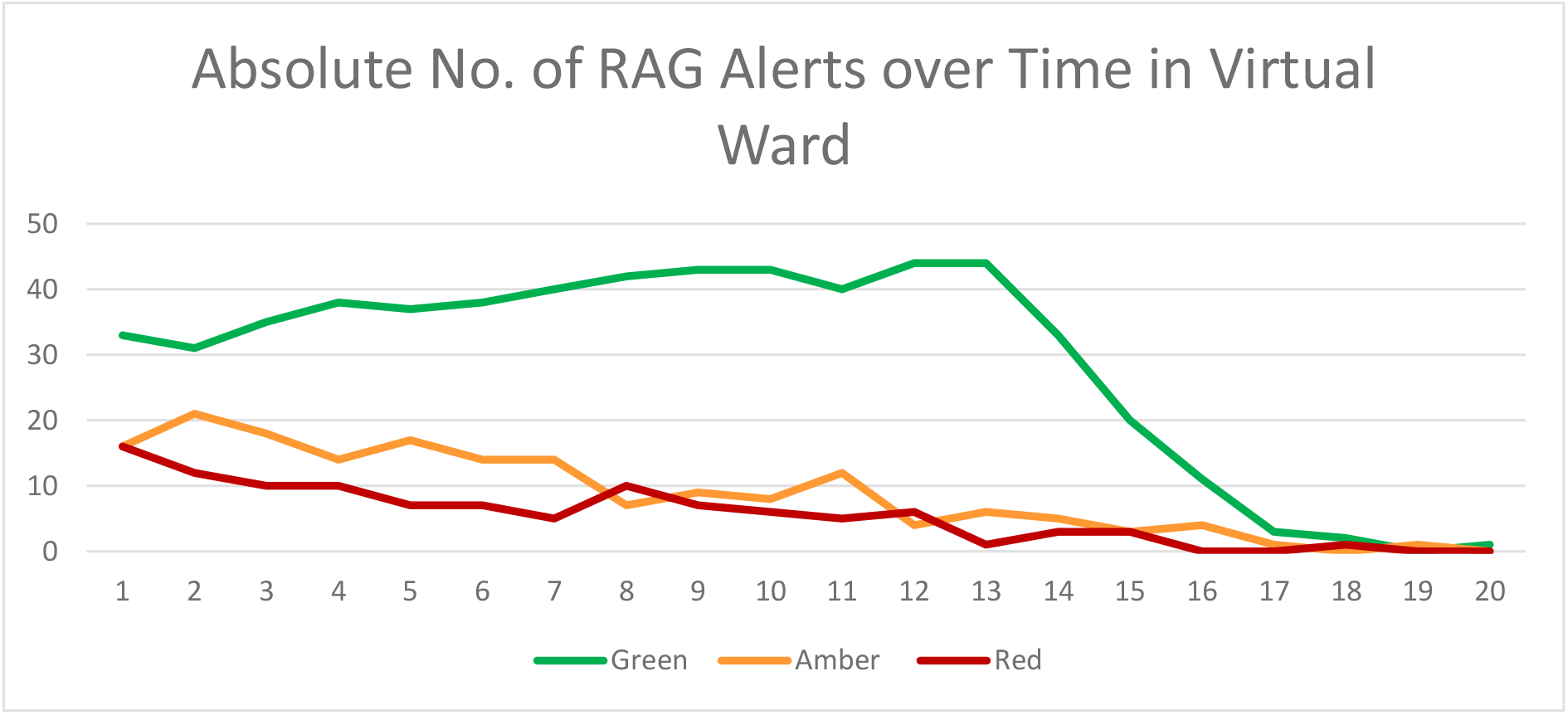
RAG Ratings Over a 20 Day Period

The mean and median length of stay in the virtual ward were 13.2 and 14 days. Patients were discharged from the virtual ward at clinical discretion. On day 15 there were still 26 patients in the virtual ward. One patient remained in the ward until day 20. Figure 2 shows the daily number of patients in the ward. 91% of patients were in the virtual ward for 9 or more days.

**Figure 2.**
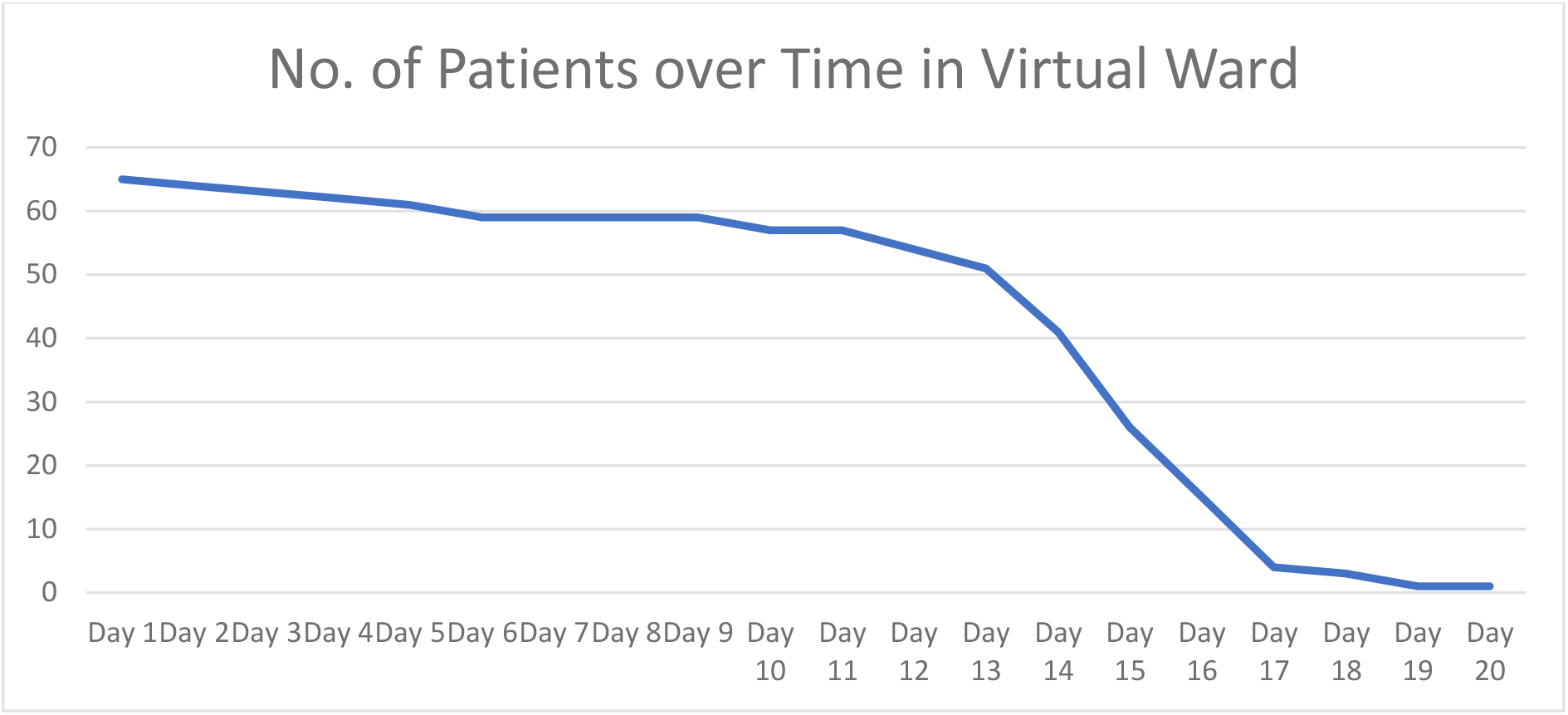
Daily No. of Patients in the Virtual Ward

There was no correlation (0.06) between the length of stay in the Glenfield respiratory wards and the length of stay in the virtual ward. The average length of stay in the hospital wards prior to the introduction of the virtual ward in November 2020 was 5.5 (+/-1.3) days^2^. The mean length of stay for those discharged into the virtual ward was 3.3 (+/-0.4) days^2^.

### Resource Use in the Virtual Ward

The total number of virtual consultations driven by a red rating was 109, 30.1% were in the first three days, see table 2. Each red rating drove two video-consultations, mean duration 27.5 minutes. These contacts were conducted by band 7 specialist respiratory nurses or physiotherapists.

**Table 2.** LPT Clinician Contact Quantity and Costs by RAG rating

| Specialist nurse calls | No. | Cost |
| --- | --- | --- |
| Reds | 109 | £2,844 |
| Ambers (week 1 only) | 114 | £2,974 |
| Greens (week 2 only) | 10 | £261 |
| Total contacts | 233 | £6,079 |

Patients with amber ratings in the first week were also contacted. There were 114 contacts that met that criterion. Ten Patients with a green rating were also contacted in the second week, where they had not been contacted before.

The total consultation costs for 65 patients over the 20 days where patients were monitored are estimated in Table 2 beneath^1^.

- The per patient cost of virtual ward patient contact was £93.52.
- The total cost of the virtual monitoring on CliniTouch Vie was £2,583, (£39.74 per patient).
- The total costs of monitoring and contacting patients were £8,662, (£133.26 per patient).
- The mean rate of re-admission was 6.2% and cost of a re-admission was estimated at £2,926^2^. The total re-admissions costs were estimated at £11,704 or £180.06 per patient.

A common adverse outcome of Covid-19 has been clotting endpoints; venous clotting has been found in 20% of patients that were hospitalised^3^. Two patients were re-admitted on day one and would likely have had their clotting event whilst in hospital under usual care and the other two patients had a clotting event on days four and five when they would likely have been discharged and then re-admitted.

### Early Discharge and Bed Days Potentially Saved

The average length of stay (ALOS) prior to and after the introduction of the virtual ward were 5.5 and 3.3 days^2^, a potential 40% reduction in bed days; p<0.001.

The mean local estimate of the cost of a bed day in a respiratory ward in The Glenfield Hospital in the month of November 2020 was estimated to be £532^2^. The implications for resource use have been summarised in Table 3 beneath.

**Table 3.** Resource Use Summary

| Elements of Resource Use | Resource Use |
| --- | --- |
| ALOS Pre-Virtual Ward Nov '20 (Days) | 5.5 |
| ALOS Discharged to Virtual Ward (Days) | 3.3 |
| No. of Patients | 65 |
| Bed Days Saved | 144.2 |
| %age Reduction in Bed Days | 40.3% |
| Bed Days Costs Averted (£) | £76,714 |
| Costs of intervention (£) | £8,662 |
| Total Resources Saved (£) | £68,052 |
| Resources Saved per Patient (£) | £1,047 |

It would appear the primary goal of increasing acute capacity was achieved. 25% of patients had red alerts on the first day in the virtual ward. It is suggested that was indicative of reduced health status and that the LPT supported discharge scheme was not taking patients that under normal circumstances would have been routinely discharged.

If the 25% of patients with a red alert on day one in the virtual ward meant that this population would have likely been retained in the hospital for an additional day, they would have cost £8,512 or 98% of the costs of the virtually supported discharge ward. There were additional red alerts in six different patients on days two and three.

The costs associated with a stay in the virtual ward were robust. For the virtual ward to be cost-neutral, it would need to have reduced one bed day for every 4.0 patients referred into it. It seemed to reduce the average number of bed days per patient per hospital admission by 2.2. The gulf between what seemed to have been achieved is 9.5 times greater than what would have been required to be cost neutral.

## Discussion

This study’s findings are limited by not having a case-matched control group and the nature of the speed of implementation. The costs of the virtual ward intervention were more certain than the costs of the status quo. The intervention was set up to reduce pressure on hospital beds as a rapid response to the pandemic. The costs have been presented transparently and outputs have been expressed as threshold values as well as point estimates. There are strengths to the implementation project, it reflected a pragmatic response to the Covid-19 pandemic and showed promising results.

Whether the supported discharge scheme was cost saving or not could be to miss the point. On January 15^th^, 48% of patients in Glenfield Hospital had been admitted with Covid-19 related disease. Creating additional capacity by minimising lengths of stay safely was considered an imperative. The implementation of the virtual ward was in tune with The Government’s agenda to “save the NHS” and funding was supported by Ageing Well. The direct costs of the virtual ward were relatively insignificant at around £133.26 per patient, which makes it a sustainable intervention, especially if patients ALOS is reduced in hospital.

No patients withdrew because of system failures or faults. If patient care will rely more heavily upon digitally supported services in the future, systems are required to be reliable enough to be enable support of clinical care. CliniTouch Vie managed a 100% record of daily sharing of patients’ data with clinicians.

One patient withdrew at their own discretion and had been green up to that point. All other patients left the virtual ward at the discretion of clinical staff.

Covid-19 has provoked a significant increase in adverse clotting outcomes^3^ and the four patients re-admitted were for that reason, one of whom died. All re-admissions were clinically reviewed by senior clinical colleagues within UHL and LPT and the inference that Covid-19 disease was the primary attributable factor for the readmission could not be ascertained. All patients were on thromboprophylaxis while in-patients and were mobile on discharge.

The LPT virtual ward was set up to better enable system capacity. A recent paper reported a 3.5 fold increase in re-admissions and 7.7 fold increase in mortality at 140 days for Covid-19 hospital discharged patients^4^. These results suggest that virtual monitoring of patients could be continued for longer, given that it is relatively inexpensive.

The reported cohort of patients registered into the service did not include patients discharged on oxygen. However, at time of print the service has been extended to those patients with the aim to ween them off this as part of their recovery from Covid-19. It is intended to report on this cohort of patients at a later date.

The digital component of the service is easily scaled up and most localities have specialist community respiratory teams. This model of care could be easily introduced in other areas, as required or expanded into additional “at risk” cohorts of patients.

The Covid-19 pandemic has accelerated a worldwide digital health revolution. The unprecedented public health challenge of providing safe care for patients and clinicians whilst also enabling increased capacity within the NHS would be difficult and expensive to sustain within traditional ways of working. Digital solutions do not replace the key role that clinicians play in the care of their patients, but they can provide them with more specific, systematic and prioritised clinical data to enable them to make quality clinical decisions.

## Data Availability

Participants in this study agreed for their anonymised data to be used for research purposes.

## Notes

### Competing Interest Statement

Swift J, OKelly NI, Barker C all work for Spirit Health Group, owners of the intellectual property associated with CliniTouch Vie. All have a potential conflict of interest. Harris Z, Woodward A and Ghosh S have no declaration of interest to declare. The virtual ward implementation project was supported by Ageing Well funding. The views and opinions expressed herein are those of the authors and do not necessarily reflect the views of Spirit Health Group, Leicestershire Partnership NHS Trust or Leicester School of Allied Health Sciences, De Montford University.

### Clinical Trial

This was a was a service evaluation, not a clinical trial

### Funding Statement

No external funding was received for the preparation of this manuscript. Swift J, O'Kelly NI and Barker C all work for the Spirit Health Group and as outlined in the competing interests section have a potential conflict of interest.

### Author Declarations

All patients gave their informed consent to the use of their anonymised data for research purposes. The evaluation proposal was submitted to Local IRB East Midlands CENTRAL Leicester and ethics was waived because it was a service evaluation. Appropriate forms have been archived.

### Summary of Updates

The data on costs was inaccurate because all specialist respiratory nurses and physiotherapists were band 7, not a mix of band 6/7. The difference made is small, but for the sake of accuracy, it was important to correct. This increased the cost per patient from 124 UK Pounds to 131 UK pounds. This had some downstream effects on some of the threshold values, which have also been altered correspondingly.

